# Estimate of the actual number of COVID-19 cases from the analysis of deaths

**DOI:** 10.1101/2020.09.21.20198416

**Authors:** Roberto Etchenique, Rodrigo Quiroga

## Abstract

Using the calculated values for the Infection Fatality Rate (IFR) of COVID-19 it is possible to estimate the prevalence of cases of infection in the city of Buenos Aires, Argentina, throughout the pandemic. The use of confirmed cases as a metric and their replacement by more reliable parameters such as death figures are discussed. The results are analyzed according to age ranges and possible sources of error in the estimates are established.

## Introduction

The number of confirmed cases of COVID is usually established in all countries through testing (usually RT-qPCR) which is done under passive (person using the health system) or active surveillance plans (search and tracing of cases through street or home operations).

There are enormous differences between the various testing systems, which leads to very different figures of positivity: from more than 3000 tests/case confirmed (New Zealand) to 1.9 test/case confirmed (Mexico). Argentina is close to the lower limit, with only 2.4 tests/cc, which corresponds to a positivity in the order of 40%. [1].

These differences make virtually useless the comparisons of case numbers between countries, between districts within a country, or even between different times within the same district or country, as the number of confirmed cases is strongly influenced by the testing strategy. While with low positive rates and efficient tracking a high percentage of actual cases can be detected, in a test saturation situation the proportion of detected cases to real cases can be very low.

For this reason it is convenient to use other methods for prevalence estimation, which do not imply the use of the number of confirmed cases, which is of little value. In countries in advanced stages of the epidemic it is useful to use the number of deaths.

Statistically, the relationship between infected and deaths has been estimated through large serology studies [2]. One of the most advanced studies was done in Spain, using seroprevalence studies, distributed by age and district [3]. By having the fraction of IgG positive in each age range, and the data on deaths due to COVID-19, which are more reliable than confirmed cases (despite possible undercounts, *vide infra*) it is possible to determine the Infection-Fatality Ratio (IFR) for each age range.

Through these rates it is possible to determine the probable number of actual cases of infection that generated a certain number of deaths. This can be done in multiple ways. A Rule-of-Thumb rule known as Garber’s Theorem was used, estimating the infections as 200 times the deaths figures. Guillermo Durán and Rodrigo Maidana calculated the proportion of cases detected in each province of the country,[4] as shown in Table 2. This estimate uses the age pyramid of each district to calculate the expected IFR if the prevalence of infections was homogeneous in all age groups.

This calculation considers that the infections are reflected in deaths 14 days after they occurred, which is a reasonable approximation, compatible with the data obtained from the Argentinean centralized data system (SISA). With this estimate it is possible to reconstruct the current value of prevalence by extrapolating the values according to the rate of increase of deaths in recent weeks.

However, there are strong reasons to expect that the prevalence will be higher in some particular age groups. Therefore, these estimates may be closer to reality if we take into account the true age pyramid of deaths, which will have in its composition not only the population age pyramid and the IFR of each range, but also the proportion in which, due to behavioral differences, each age range is susceptible to infection. This preliminary work addresses the case of the Autonomous City of Buenos Aires (CABA) as an example for the determination of infections by the IFR method of cases of death.

## Methods and Results

The basis of the proposed calculation is to use the reciprocal function of the IFR, which indicates the number of infections corresponding to each death. Although this can be done directly with the data of each range, we chose to fit a smooth log-polynomial curve (to reduce the noise of the determination) to the IFR values obtained from the Spanish study. To do this, equation (1) was fitted to the centers of the ranges referred to in Table 1.

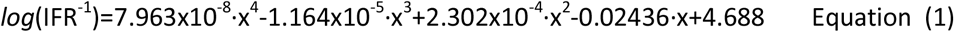

**Table 1:** Results of the seroprevalence study in Spain (data compiled in by @gummybear737) https://twitter.com/gummibear737/status/1262453620735385605

| Age | Population | Incidence as per AB Testing | Total Infected | Cases Recorded (Data - May 5th) | % Cases Recorded | % Mild or Asymptomatic | Deaths | IFR |
| --- | --- | --- | --- | --- | --- | --- | --- | --- |
| 0-9 | 4,340,500 | 2.61% | 113,287 | 871 | 0.77% | 99.23% | 2 | 0.002% |
| 10-19 | 4,682,400 | 3.85% | 180,344 | 1,331 | 0.74% | 99.26% | 5 | 0.003% |
| 20-29 | 4,652,000 | 4.64% | 216,061 | 12,067 | 5.58% | 94.42% | 22 | 0.010% |
| 30-39 | 6,158,200 | 4.25% | 261,964 | 20,386 | 7.78% | 92.22% | 57 | 0.022% |
| 40-49 | 7,935,300 | 5.50% | 436,677 | 31,658 | 7.25% | 92.75% | 183 | 0.042% |
| 50-59 | 6,944,600 | 5.95% | 413,128 | 38,792 | 9.39% | 90.61% | 567 | 0.137% |
| 60-69 | 5,200,500 | 6.03% | 313,678 | 31,805 | 10.14% | 89.86% | 1526 | 0.486% |
| 70-79 | 3,921,700 | 6.56% | 257,283 | 30,632 | 11.91% | 88.09% | 4273 | 1.661% |
| 80-89 | 2,318,300 | 5.30% | 122,943 | 34,032 | 27.68% | 72.32% | 7117 | 5.789% |
| 90+ | 583,100 | 5.80% | 33,820 | 15,934 | 47.11% | 52.89% | 3496 | 10.337% |

**Table 2:** Prevalence calculation in Argentine provinces using the IFR in table 1

| Districts | Theoretical IFR | Confirmed cases incidence | Infected / Cases ratio | Estimated infected population at 28-Aug | Projected infected population at 11-Sep |
| --- | --- | --- | --- | --- | --- |
| **Argentina** | **0.33%** | **0.91%** | **8.1** | **7.44%** | **9.57%** |
| Buenos Aires | 0.34% | 1.48% | 7.6 | 11.22% | 13.88% |
| Córdoba | 0.35% | 0.21% | 6.8 | 1.42% | 2.43% |
| CABA | 0.52% | 3.05% | 5.5 | 16.66% | 19.20% |
| Catamarca | 0.28% | 0.02% | 0.0 | 0.00% | 0.00% |
| Chaco | 0.23% | 0.44% | 19.3 | 8.47% | 10.47% |
| Chubut | 0.25% | 0.15% | 4.3 | 0.63% | 1.02% |
| Corrientes | 0.27% | 0.03% | 3.5 | 0.10% | 0.19% |
| Entre Ríos | 0.34% | 0.23% | 8.0 | 1.87% | 2.88% |
| Formosa | 0.25% | 0.01% | 4.9 | 0.07% | 0.08% |
| Jujuy | 0.25% | 1.10% | 13.0 | 14.34% | 19.72% |
| La Pampa | 0.37% | 0.06% | 4.1 | 0.22% | 0.36% |
| La Rioja | 0.25% | 0.38% | 20.4 | 7.66% | 13.81% |
| Mendoza | 0.33% | 0.36% | 8.3 | 2.98% | 5.47% |
| Misiones | 0.22% | 0.00% | 16.2 | 0.07% | 0.08% |
| Neuquén | 0.25% | 0.43% | 10.4 | 4.53% | 7.00% |
| Río Negro | 0.29% | 0.79% | 13.6 | 10.67% | 15.18% |
| Salta | 0.24% | 0.23% | 11.1 | 2.52% | 4.81% |
| San Juan | 0.28% | 0.03% | 12.3 | 0.32% | 0.63% |
| San Luis | 0.30% | 0.01% | 0.0 | 0.00% | 0.00% |
| Santa Cruz | 0.18% | 0.47% | 7.3 | 3.44% | 5.47% |
| Santa Fe | 0.38% | 0.21% | 6.8 | 1.40% | 3.20% |
| Santiago del Estero | 0.25% | 0.08% | 11.0 | 0.92% | 1.85% |
| Tierra del Fuego | 0.16% | 1.18% | 13.2 | 15.58% | 20.06% |
| Tucumán | 0.27% | 0.12% | 3.2 | 0.40% | 1.12% |
| **AMBA** | **0.32%** | **2.03%** | **8.4** | **17.08%** | **20.47%** |

Figure 1 depicts the obtained result:

**Figure 1.**
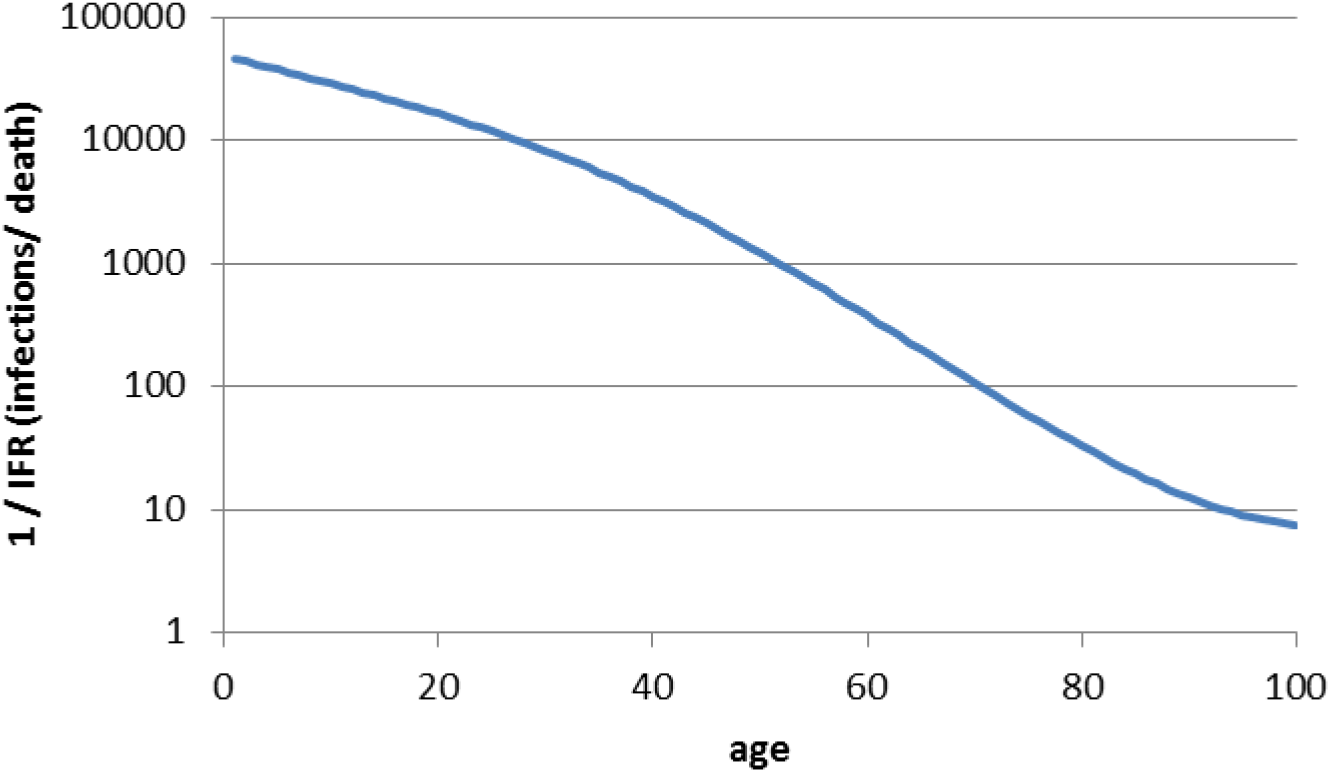
reciprocal IFR function representing the number of infections required for each subsequent death.

Figure 2 shows the result of the transformation of each death in a number of cases according to the curve in Figure 1 for CABA. The high peaks that are seen are the product of the few deaths that occurred in individuals of low age, which are interpreted as the product of a large number of point infections with a delay of 11 days in their death with respect to the date of infection. While these curves can be smoothed out, they do not reflect the likely dynamics of the deaths. It is important to note that the final drop does not correspond to a real trend but rather to the undercounting of deaths by two factors: 1) the infection-death time is very variable and 2) many districts are slow to register a large proportion of deaths in the system.

**Figure 2:**
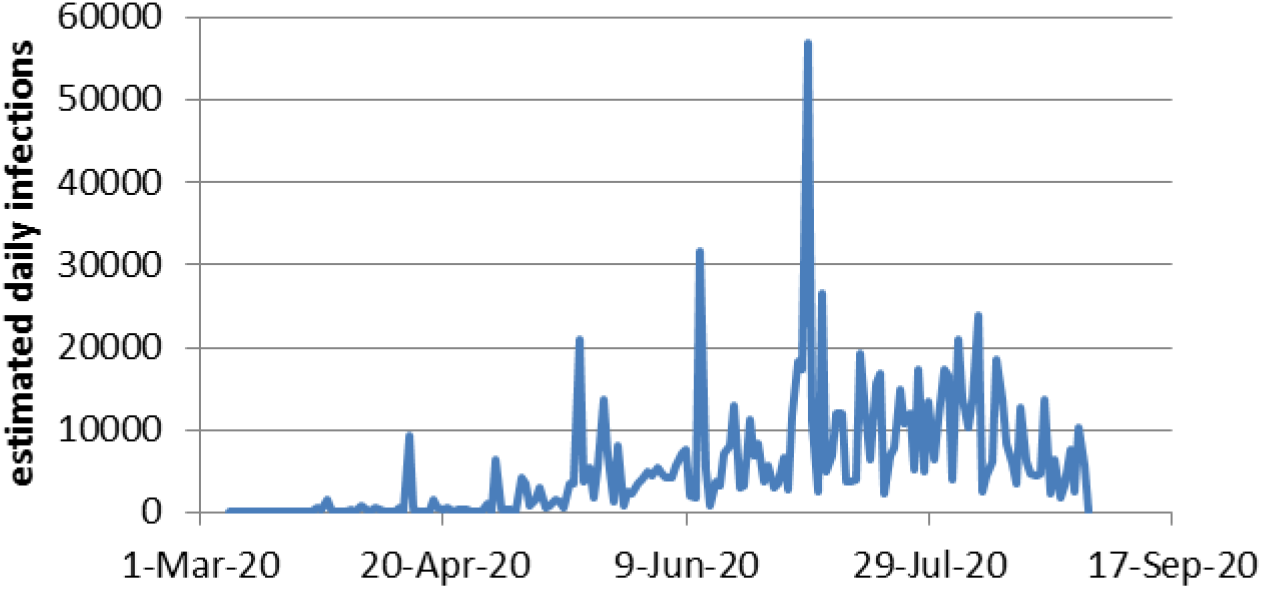
Transformation of each CABA death into a number of infections 11 days earlier through the curve in Figure 1. N_inf_(t-11) = N_dead_(t)/IFR(age)

Thus, each death registered in the Argentinean Integrated Health Information System will have a previous number of cases 1/IFR. Although this number of estimated actual infections can be entered in a single day, in that case the temporary values obtained will be noisy, especially for the age ranges of lower IFR.

To eliminate these problems, an infection-to-death function obtained from the experimental data was used. Figure 3 shows the histogram from the recorded date of symptom onset to the date of death. The data were fitted with the following empirical expression:

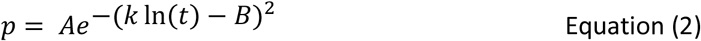

**Figure 3:**
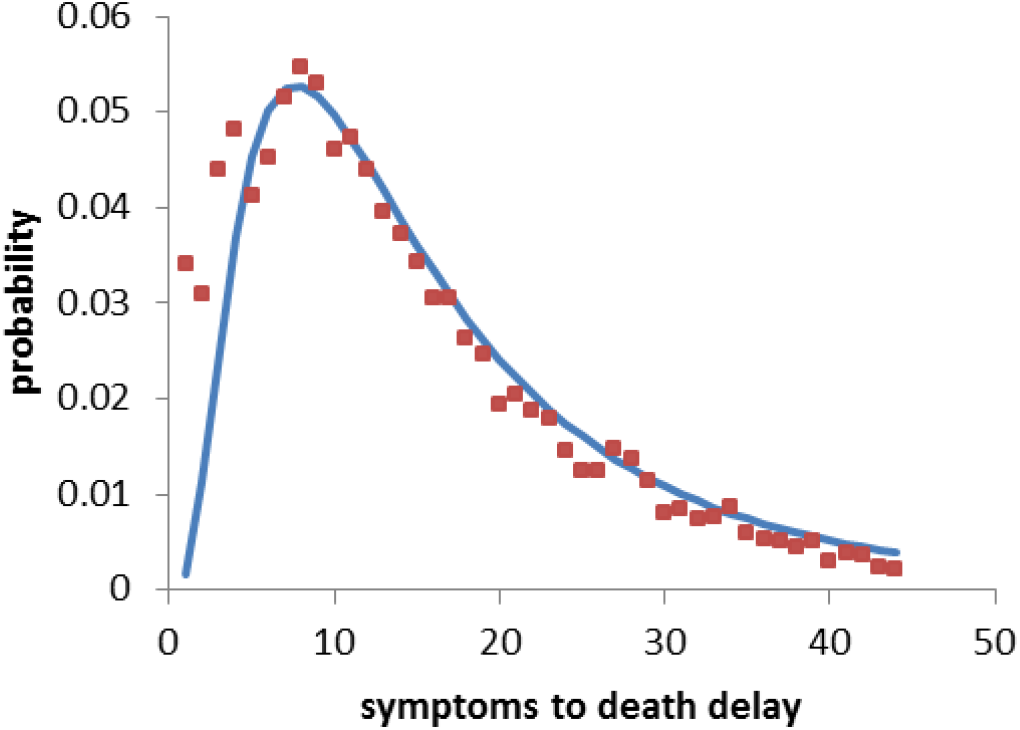
Days between onset of symptoms and date of death according to SISA for the Buenos Aires metropolitan area. Fitted parameters: A= 5.28×10^−2^; B=1.883; k=0.924

Once this statistical dependency is established, each death will be counted as 1/IFR number of cases distributed in the previous 44 days according to the previous curve.

A typical result of this procedure is shown in Figure 4.

**Figure 4.**
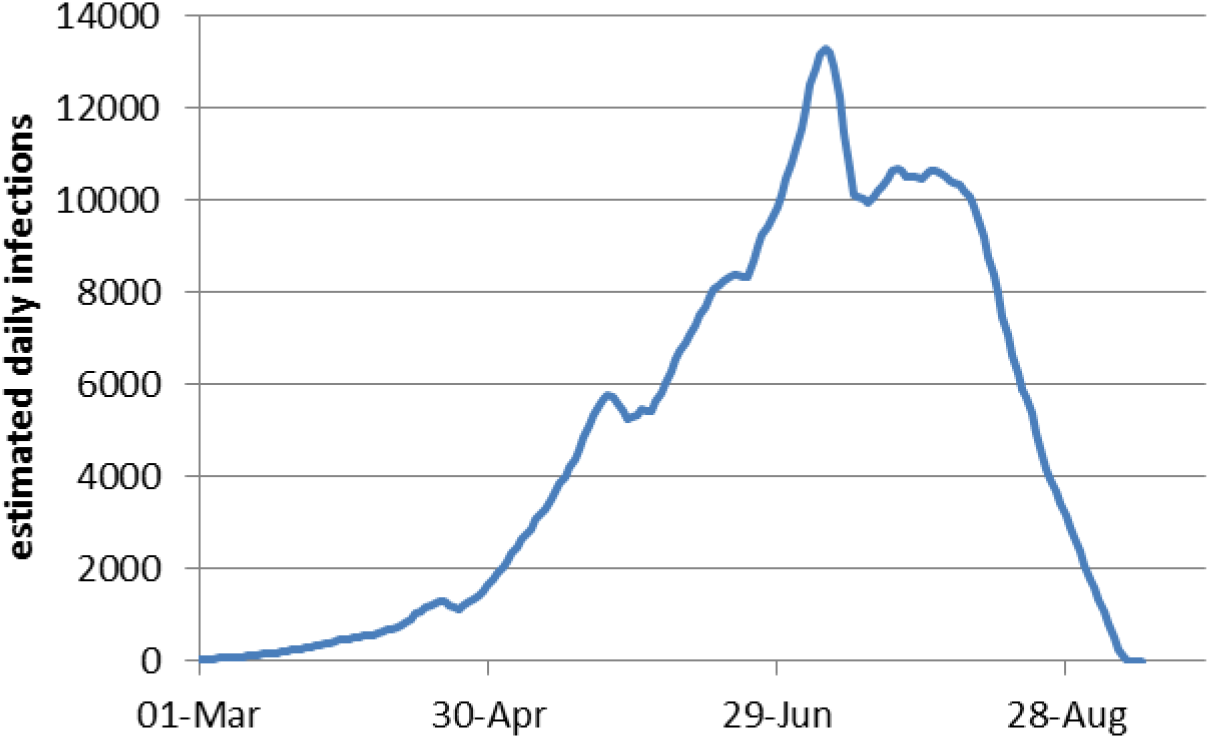
Estimate of daily infections in CABA according to the transformation of each death into a series of infections that follow the probability of occurrence in Figure 3.

The local maxima observed in mid-May and June correspond to events of death of young people with very low IFR. If restricted to those over 30 years of age, the dependency takes on the appearance of Figure 5.

**Figure 5:**
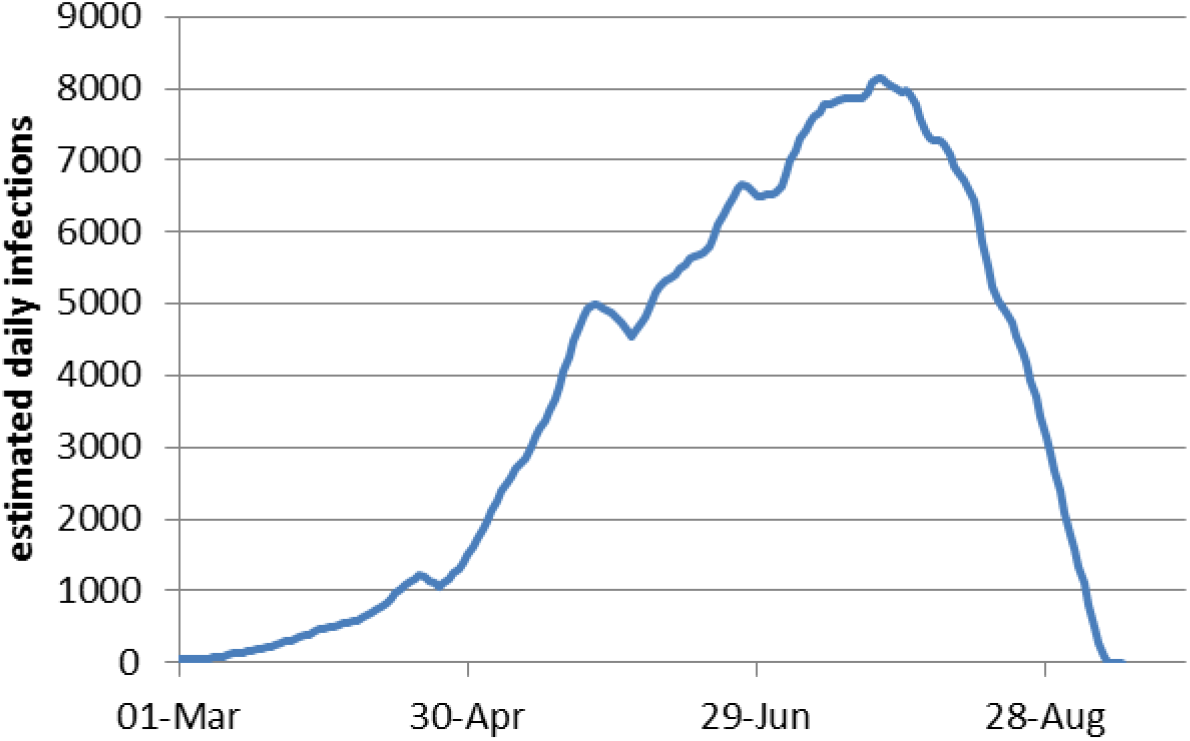
Estimate of daily infections in CABA according to the transformation of each death into a series of infections that follow the probability of occurrence in Figure 3.

The decrease observed at dates at the end of the graph, close to that of the used dataset is again the result of data censoring. Many cases of infection that will end in death are not yet counted, and a proportion of cases that have already died have not been loaded into the system. To solve this problem, a normalizing function was built by using the data recorded on 1/8/2020 and comparing it with the data from 11/9/2020, the date on which all the July data are supposed to be stable. This function is defined by the following empirical expression

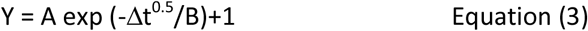

in which Δt is the temporal distance in days between the last date of the dataset and the date of estimated cases of infection, A = 53.84 and B=1.29, values obtained by fitting the old data to the new one using the transformation D_old_·Y = D_new_ The result of applying this transformation is shown in Figure 6.

**Figure 6.**
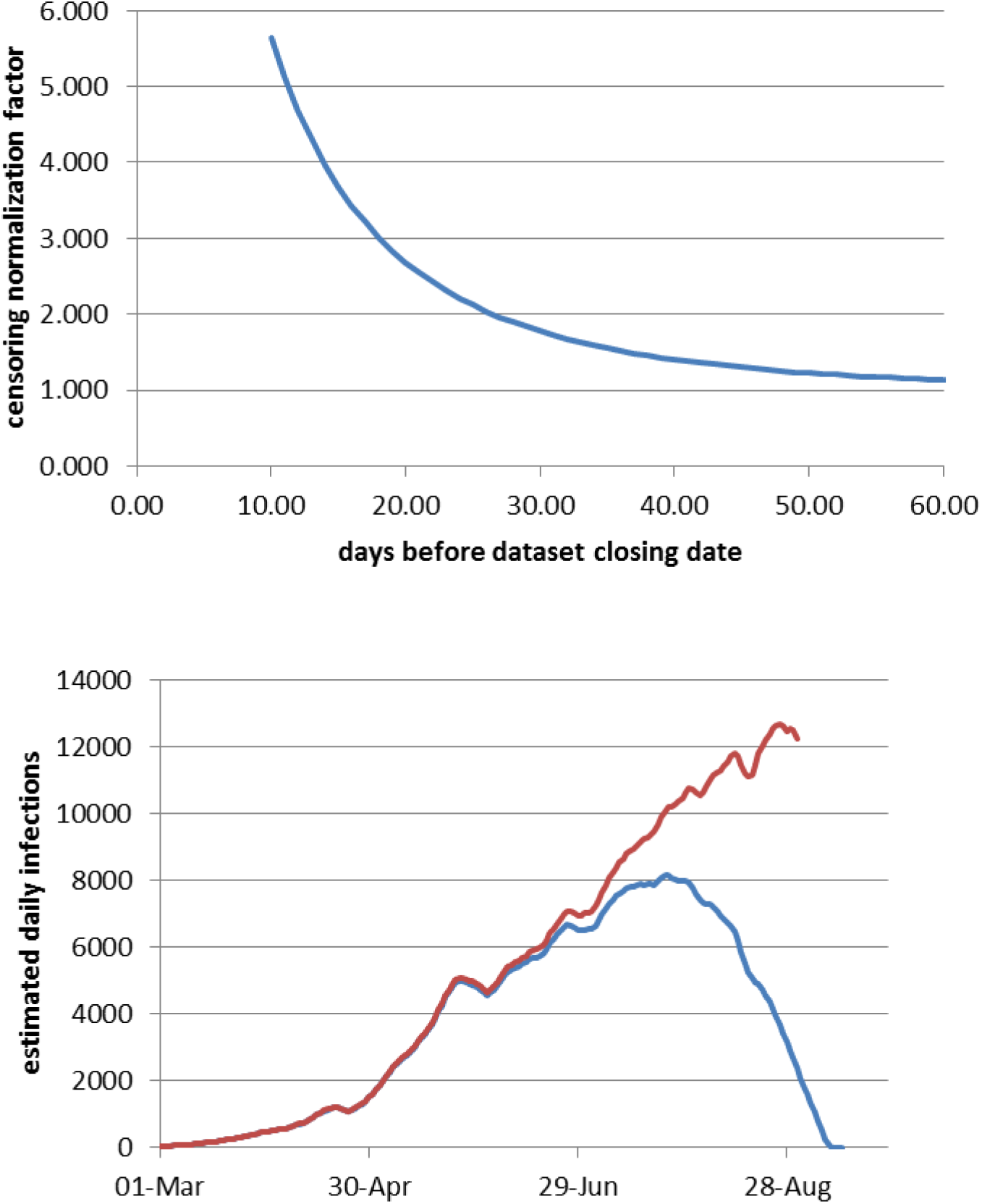
Estimated daily CABA infections for those over 30 years old. Above: censoring normalization function. Below: Blue: raw case curve. Red: corrected case curve with the normalization function.

Since the data have been obtained with IFR as a function of age, it is possible to determine the dynamics of each age range individually. The result, once normalized, is shown in Figure 7.

**Figure 7.**
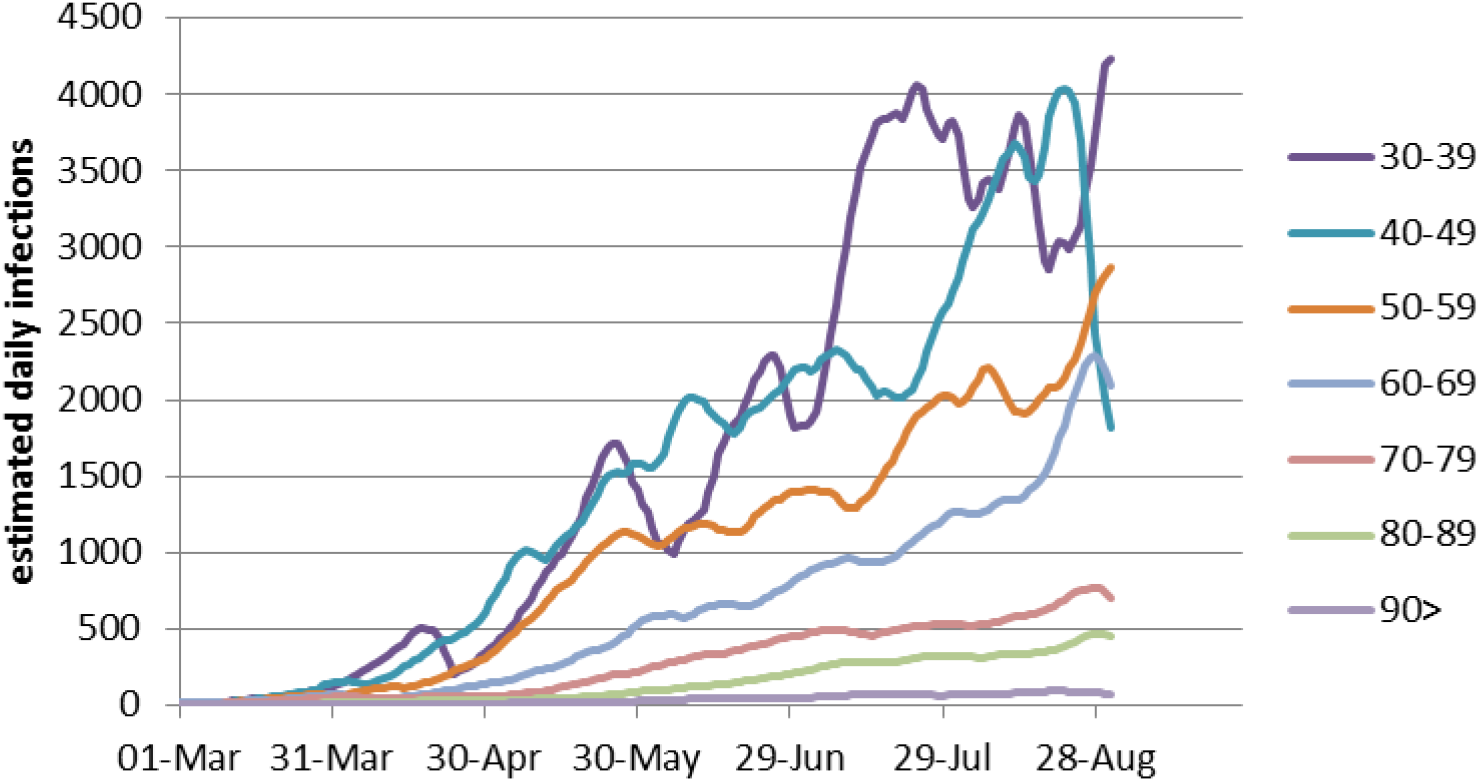
Estimated daily infections in CABA by age range. The ranges from 0 to 29 are not shown due to the noise due to a reduced number of deaths.

The time integration of the curves in Figure 7 corresponds to the cumulative prevalence for the city of Buenos Aires and can be seen in Figure 8. The estimated prevalence as of August 31, 2020 is shown in Figure 9.

**Figure 8:**
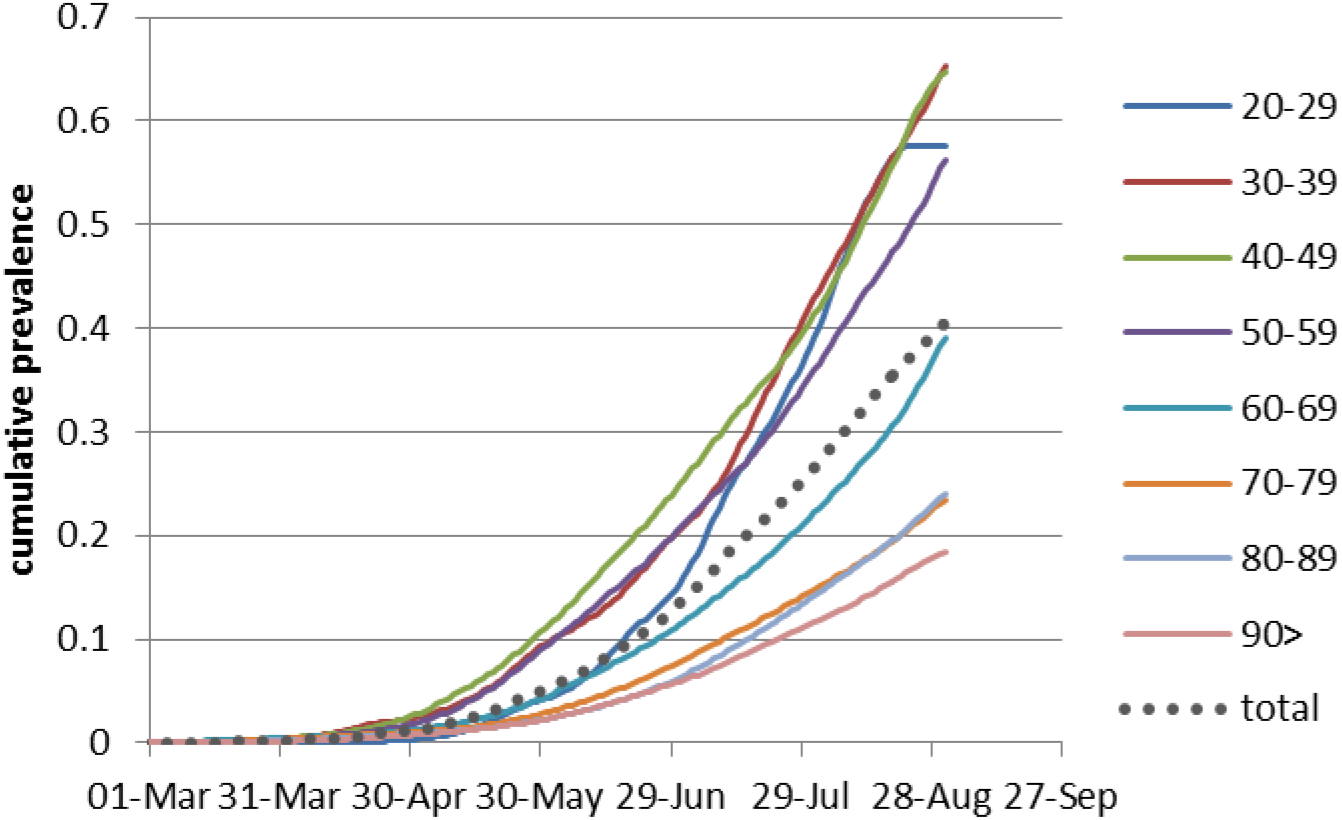
Evolution of prevalence for various age ranges in CABA

**Figure 9.**
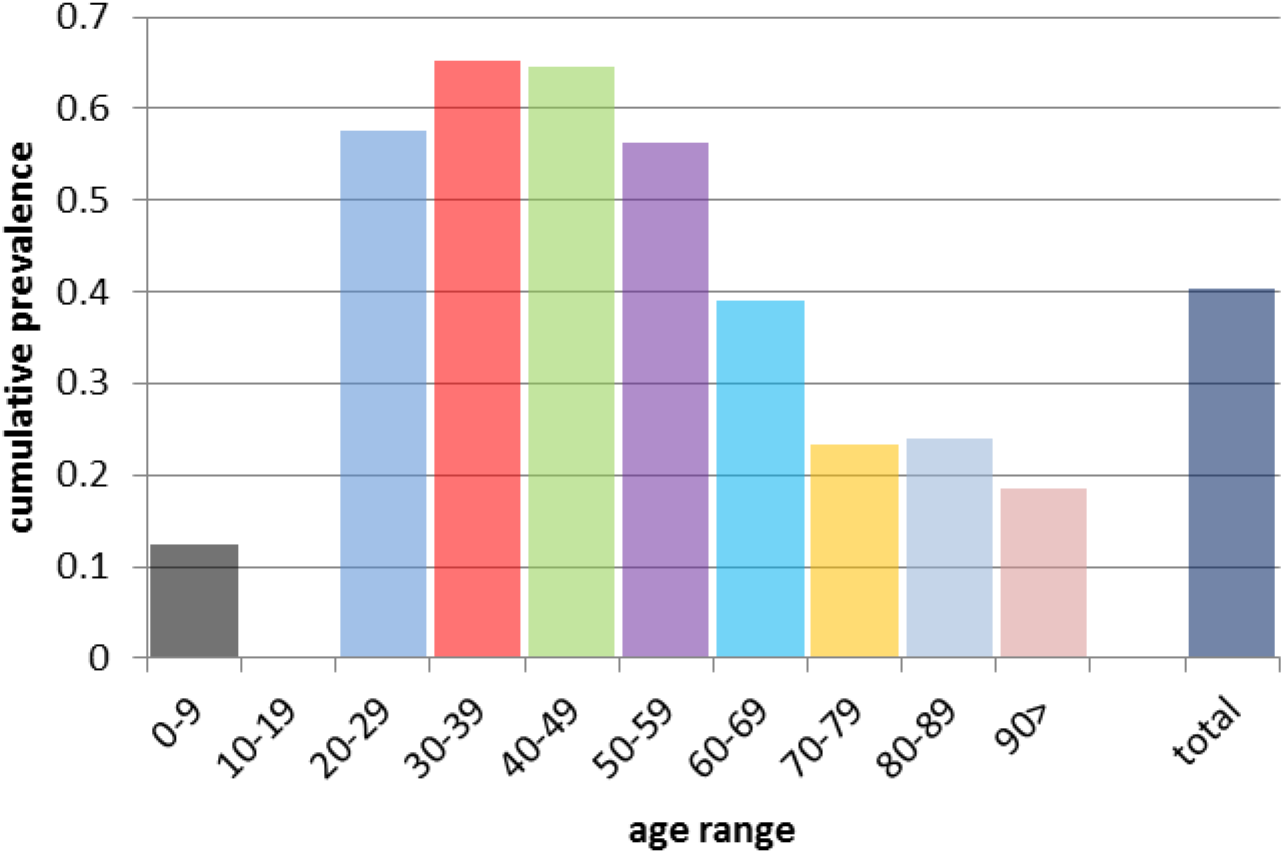
Cumulative prevalence at 8/31/2020 in the Autonomous City of Buenos Aires according to age range, estimated from IFR values.

## Discussion

The use of “confirmed cases” as a widespread metric throughout the world, especially in the media and social networks has many implications not usually considered. One of them is the factual impossibility of comparing situations in 2 different regions, countries, districts, and even within the same area at two different times. This is because the relationship between “confirmed cases” and actual cases of SARS-COV-2 infection is extremely uncertain, and depends heavily on the testing policy and strategy and the capacity of the system to manage large numbers of infections in a timely manner. Only a tiny minority of countries have achieved a testing system that roughly reflects the actual number of infections, which is particularly difficult since many of them are asymptomatic, with estimates varied from 15% to 80% of those infected. [6,7] The terminology frequently used in the media, calling confirmed cases “contagions”, suggests that this number of reported cases actually corresponds to those infected. However, it is common to find whole countries where the number of confirmed cases can be estimated to be 20-30 times lower than the actual number (e.g., Belgium) or cities where the ratio was 50 to 1 (London). This effect is exacerbated when in a situation of mass transmission and system saturation.

The Mandatory Social Preventive Isolation (ASPO, from its initials in Spanish) was nationwide in Argentine at March 20th, and was effectively complied with in all districts for approximately 2 to 3 weeks. From that moment on, different districts carried out different policies in the management of the pandemic.

In particular, the Autonomous City of Buenos Aires had an openness policy from the beginning, taking to the extreme the original idea of “flattening the curve” of infections (aka mitigation, as contrary to suppression) so that the population would be infected at a low rate in order to prevent the collapse of the health system, but not adopting active policies that would try to eliminate or reduce the number of daily infections. Additionally, the policy of control and sanitary isolation of those who arrived in Argentina in repatriation flights from countries with very high viral circulation was not very effective.

They were isolated in hotels, with subsequent swabbing and return home in case of negative results. The high rate of false negatives in PCR during the first days after infection [8], and the possible subsequent failure to comply with the mandatory isolation in homes, may have produced a continuous influx of infected during the months of March to June, which may explain the inability to reduce cases to very low levels as happened in the rest of the Argentine provinces.

After 4 months of rising daily confirmed cases, the dynamics of cases in CABA began a series of rises, stabilizations and gentle declines that remained around the 1000 confirmed cases per day on average. This trend is shown in Figure 8.

**Figure 8:**
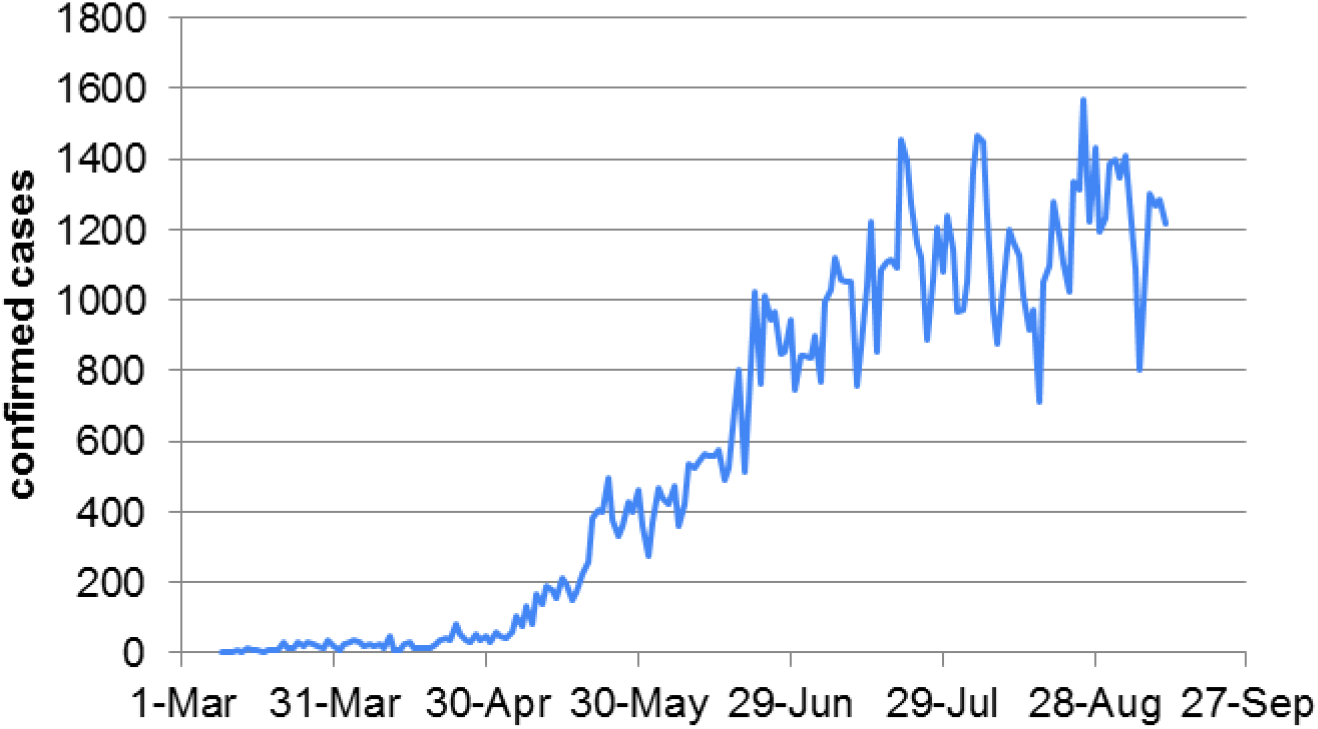
confirmed cases at CABA by date of report. (Source: Jorge Aliaga’s database based on official data from the Ministry of Health)

However, the positivity of the tests in the order of 40 to 50% on most days indicates that the actual number of cases per day should be much higher than those reported. For this purpose, the number of deaths through the IFR can be used, obtaining the dynamics shown in Figure 6 and following.

Through this analysis, it can be seen that the curve of cases estimated from the IFR is approximately 10 times higher than the curve of confirmed cases, indicating that 90% of cases go undetected. On the other hand, the shape of the curve of confirmed cases shows a saturation effect that is not observed in the curve of infections estimated from the IFR, suggesting that the limit imposed by the testing system is responsible for the high positivity and the subsequent loss of cases. This behavior can be seen in Figure 9.

**Figure 9.**
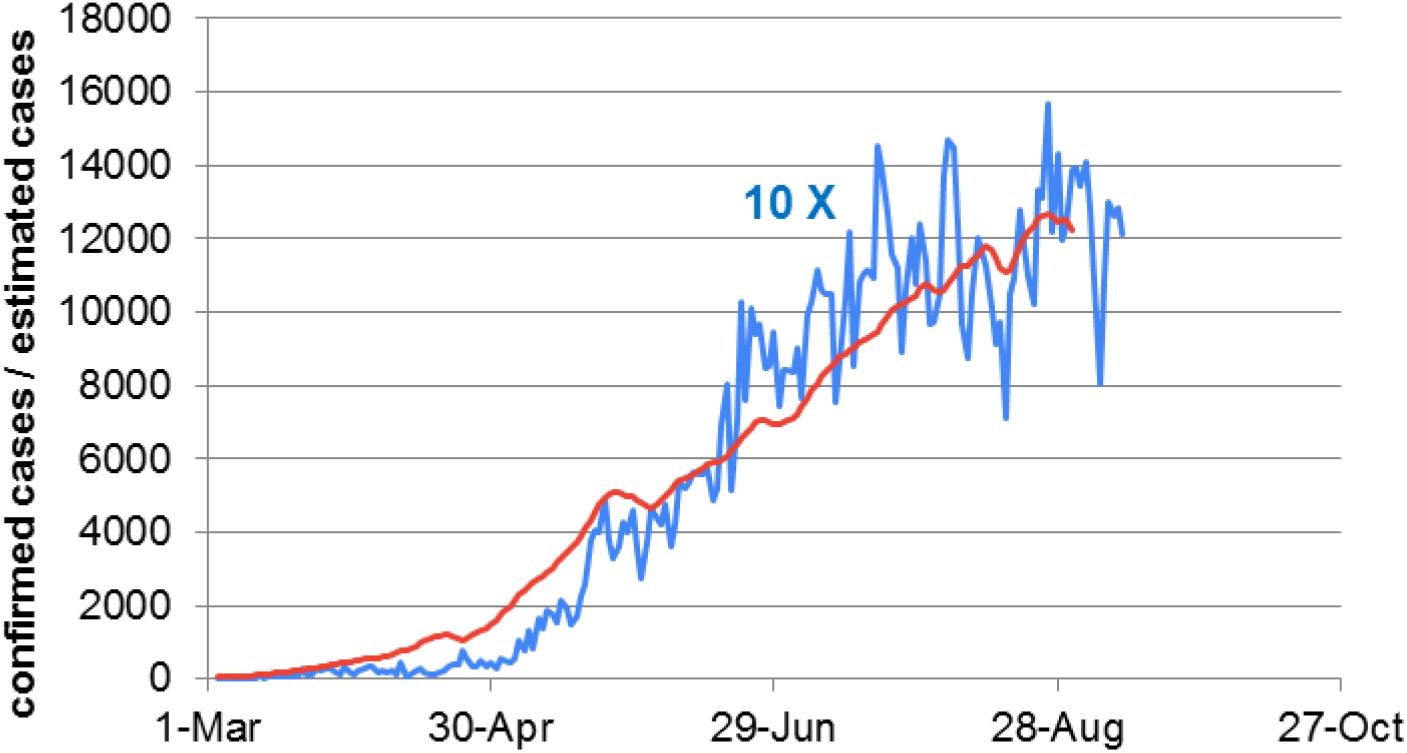
Comparison of the curve of confirmed cases by date of official confirmation (blue) with the dynamics of actual infections obtained from the analysis of deaths through IFR (red). The scale corresponds to the IFR curve. The confirmed cases curve is magnified 10 times to facilitate the reading of the graph.

This ratio of approximately 10 to 1 has important consequences when analyzing the situation of the City. The most important is that the number of confirmed cases accumulated at the end of August corresponds to approximately 3.3% of the total population. This number sets a lower limit for cumulative prevalence. In contrast, the integration of the cases estimated by IFR corresponds to 40% of the total population of CABA, which would indicate a completely different situation. The analysis by age range of this 40% indicates that while in the range of 20 to 60 years the contagion would reach about 50% of the population, the prevalence in senior citizens is much lower. This may be due to fewer daily social contacts by seniors [9], the greater caution they have taken to protect themselves from the virus (fright immunity, “inmunidad de cagazo” in original Argentinean slang) and/or the fact that people in their 20s and 30s make up the majority of active workers. There were no specific health measures to reduce the exposure of the older population, except for some preventive and early detection measures in geriatric hospitals. On the other hand, despite the noise inherent in the low IFRs of these age ranges, the curves corresponding to individuals aged 20-49 years suggest stagnation, perhaps due to the higher rate of previous infection. This trend is not clearly seen in the other ranges.

If this is indeed the case, this situation poses an imminent danger. By lowering the number of daily cases due to the reduction of the susceptible population through infection in conjunction with reduced mobility and social connectivity through current health regulations, the recovery of confidence and “back to normality” of the elderly in an environment of high viral circulation could lead to a significant increase in the death rate without a large prior increase in the number of cases, given the high IFR in adults over 60.

However, the case growth curves shown in Figure 7 suggest that an overall prevalence of 40%, with peaks of 60% in the most mobile sector of society may be the result of an underestimation of the IFR. Assuming that individuals are immune to reinfection (at least in short time), for the values of the expected and estimated R reproduction number in conditions of social distancing, and confirmed by the low slope of the infection curve, a cumulative prevalence of 40% should be accompanied by a clear lowering of cases, beyond the sanitary measures provided, which is not observed, although it is glimpsed in the last week analyzed.

A possible source of error is the undercounting of deaths in Spain, which is known given the figures of excess death over previous years. While the confirmed COVID deaths as of June 21 are 22,000, the excess deaths for that date total 48,000. Some of these deaths may be due to the collapse of the health system, but many others probably correspond to unrecorded cases of COVID-19. This could raise the IFR values to twice their reported value. If this were the case, the cumulative prevalence values in CABA could be around 20%. The only prevalence study made public in CABA was conducted between May 15 and July 18, and indicated about 9% for the estimated dates [10]. However, this low prevalence would indicate that the IFR values in Argentina would be much higher than the values reported in Europe. On the other hand, the unreported deaths in Spain are considered to have occurred primarily among older people, many of whom lived in nursing homes. This would imply that only the IFRs in these ranges would be higher, further lowering the calculated prevalence.

Another possible cause of error due to excess prevalence could be the socioeconomic conditions of CABA, which on average could be worse than the Spanish average weighted by the regions where there were more infections. This situation could be aggravated by factors related to the health system. However, although this could be the reality in other regions of Argentina, the City of Buenos Aires has a GDP/capita of values close to those of Europe, although much more unevenly distributed. This situation could distort the age distribution of the IFR.

Of these possible causes of error, the underestimation of the IFRs in Spain due to the undercounting of deaths by COVID is more plausible. More recent estimates of IFR values [3] using the figures for excess deaths during the initial period of the pandemic in Spain tend to confirm this possibility. Although these new values are erratic in the lower age ranges, due to a significant amount of excess deaths which do not necessarily have to be attributed to COVID-19, a log-linear extrapolation to these low ranges allows estimating these IFR values. On the other hand, this new work does not differentiate between the 80-89 and 90+ age ranges, an aspect that is particularly important due to the large difference in IFR observed between the 80-89 and 90+ age groups. Figure 10 shows the IFR values corresponding to both estimates.

**Figure 10.**
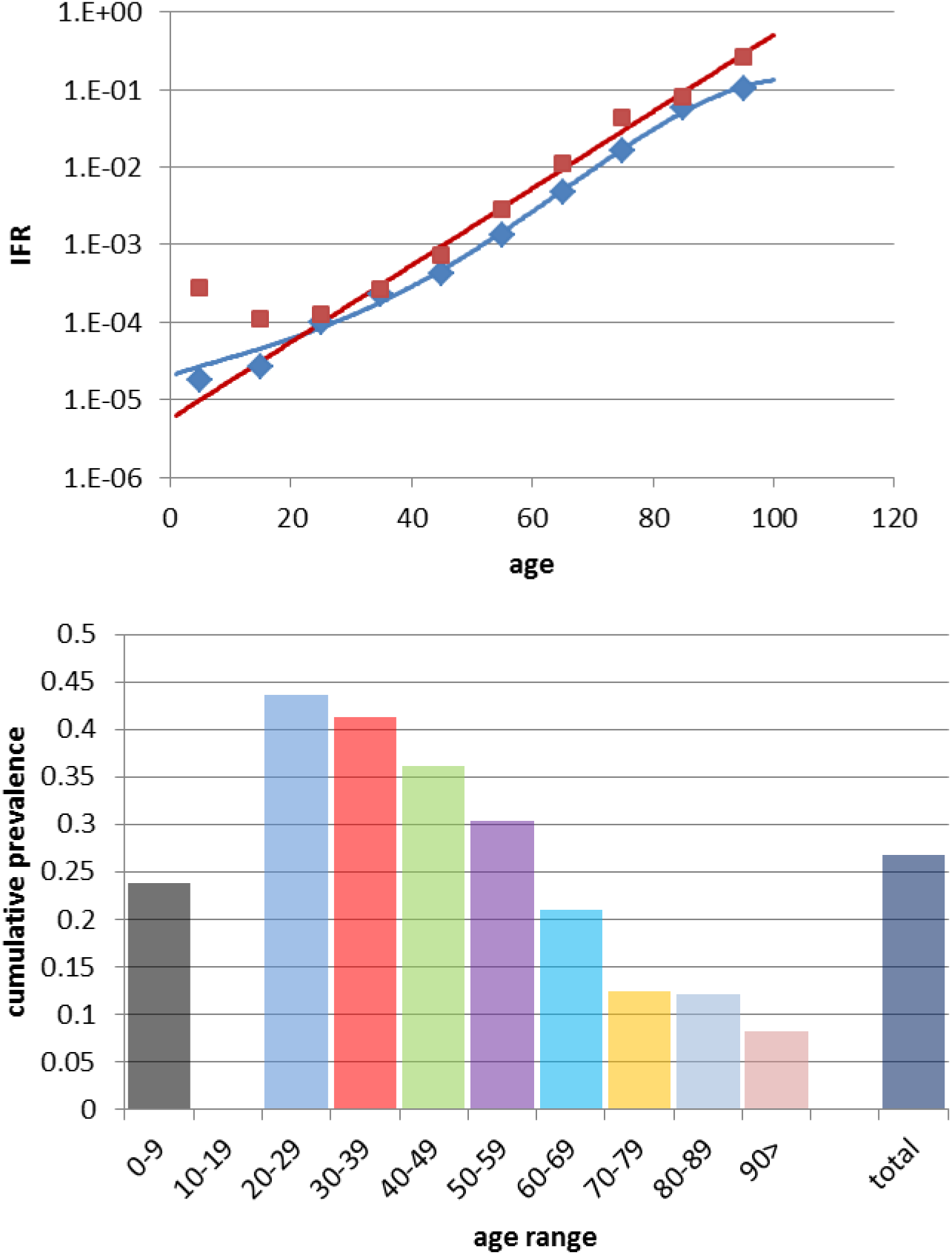
a) IFR originally calculated from confirmed COVID-19 data deaths from Spain. Diamonds: Raw IFR by age range. Blue line: log-polynomial interpolation. Squares: New IFR using excess of deaths as estimator. Red line: log-linear interpolation, without taking into account the lowest age ranges. b) Accumulated prevalence at 31/8/2020 in the Autonomous City of Buenos Aires according to age range, estimated from IFR values using excess deaths values.

Using the loglinear expression for the corrected IFR, the prevalence values of Figure 11b are obtained, more in accordance with the observed slope in cases and death curves. Other considerations will be included in future versions of this work and it will be extended to other districts where death analysis is possible.

## Data Availability

All used data is public.

## Acknowledgments

RE and RQ are members of CONICET. Fruitful discussions with G.Duran, R.Castro, J.Aliaga and J.Tiffenberg are kindly acknowledged.

